# Substance Use and Pre-Hospital Crash Injury Severity Among U.S. Older Adults: A Five-Year National Cross-sectional Study

**DOI:** 10.1101/2022.06.14.22276313

**Authors:** Oluwaseun Adeyemi, Marko Bukur, Cherisse Berry, Charles DiMaggio, Corita Grudzen, Abidemi Adenikinju, Allison Cuthel, Jean-Baptiste Bouillon Minois, Omotola Akinsola, Alison Moore, Joshua Chodosh

**Affiliations:** Ronald O Perelman Department of Emergency Medicine, New York University Grossman School of Medicine, New York, USA; Department of Surgery, New York University Grossman School of Medicine, New York, USA; Department of Population Health, New York University Grossman School of Medicine, New York, USA; Department of Medicine, Memorial Sloan Kettering Cancer Center, New York, USA; Department of Orthopedics, Mayo Clinic, Minnesota, USA; Emergency Department, CHU Clermont-Ferrand, Clermont-Ferrand, France; Department of Social Work, Minnesota State University, Minnesota, USA; Department of Medicine, University of California San Diego, California, USA; Department of Medicine, New York University School of Medicine, New York, NY, USA; Medicine Service, Veterans Affairs New York Harbor Healthcare System, New York, NY, USA

**Keywords:** substance use, crash injury, older adults, rurality/urbanicity, fatal injuries

## Abstract

**Background:** Alcohol and drug use (substance use) is a risk factor for crash involvement.

**Objectives:** To assess the association between substance use and crash injury severity among older adults and how the relationship differs by rurality/urbanicity.

**Methods:** We pooled 2017 – 2021 cross-sectional data from the United States National Emergency Medical Service (EMS) Information System. We measured injury severity (low acuity, emergent, critical, and fatal) predicted by substance use, defined as self-reported or officer-reported alcohol and/or drug use. We controlled for age, sex, race/ethnicity, road user type, anatomical injured region, location (scene) of the injury, rurality/urbanicity, time of the day, and EMS response time. We performed a partial proportional ordinal logistic regression and reported the odds of worse injury outcomes (emergent, critical, and fatal injuries) compared to low acuity injuries, and the predicted probabilities by rurality/urbanicity.

**Results:** Our sample consisted of 253,933 older adults (65 years and older) road users. Approximately 67%, 25%, 6%, and 1% sustained low acuity, emergent, critical, and fatal injuries, respectively. Substance use was reported in approximately 3% of the population, and this proportion did not significantly differ by rurality/urbanicity. After controlling for patient, crash, and injury characteristics, substance use was associated with 35% increased odds of worse injury severity. Compared to urban areas, the predicted probabilities of emergent, critical, and fatal injuries were higher in rural and suburban areas.

**Conclusion:** Substance use is associated with worse older adult crash injury severity and the injury severity is higher in rural and suburban areas compared to urban areas.

## Introduction

Every day in the United States (U.S.), approximately 700 older adults sustain crash injuries with varying degrees of severity[1]. As of 2020, there were over 44 million licensed older adult drivers in the U.S. - a 68% increase compared to two decades ago [1]. These older adult drivers are at increased crash risk due to low visual acuity, poor peripheral vision, presence of other eye diseases, hearing loss, decline in motor skills, and other environmental road conditions such as nighttime driving [2,3]. However, crash injuries involving older adults extend beyond being car occupants but include pedestrians, riders of bicycles and tricycles, and bus or truck occupants. While motor vehicular crashes account for about 55% of crash injuries among older adults,[4,5] pedestrian crash rates (secondary to motor vehicle use) have also been on the rise, increasing from 40.7 to 45.0 per 100,000 population between 2009 and 2019 [6].

Further increasing the risk of crash involvement and injury among older adults is alcohol and drug use (collectively referred to as substance use) [7–10]. It is estimated that approximately 38,000 older adults receive opioid prescriptions every day, one out of every eight older adults takes alcohol daily, and a smaller proportion report daily use of marijuana and cocaine [11,12]. Across all age groups, substance use while driving is associated with 45% increased odds of adverse crash outcomes and increased risk of pre-hospital crash fatality [13,14]. Alcohol is associated with two to seven folds increased odds of crash involvement in a crash,[15–17] and a 15-fold increased odds of severe injury [16]. Marijuana, opioids, narcotics, stimulants, and depressants are associated with two to six folds increased crash risks and odds of fatal crash injuries [18–23].

Although crash injuries are preventable, rapid provision of care can improve injury outcomes among older adults. The rural-urban disparity in Emergency Medical Service (EMS) response,[24,25] may disproportionately predispose older adults with crash injuries to worse injury severity compared to older adults with similar injuries in urban areas [26]. Earlier studies have reported that, while fatal injuries occur more in rural areas, minor and serious injuries occur more in urban areas [14,27,28]. Additionally, substance use differs across rurality/urbanicity. While urban areas have a higher incidence of hallucinogens, cocaine, marijuana, and other illicit drug use, rural areas have a higher incidence of alcohol and opioid misuse [29].

It is unknown to what extent substance use is associated with injury severity among older adults. Additionally, it is not known how the relationship between substance use and crash injury severity among older adults differs across rural and urban areas. Identifying these regional differences may inform policies on safe driving, road infrastructural design, and targeted behavioral interventions for older adults. Assessing the risk of crash injury severity among older adults is important due to the increasing older US adult population,[30,31] and older licensed drivers [1]. This study, therefore, aims to assess the relationship between substance use and crash injury severity among older adults and the rural-urban differences that further define this problem.

## Methods

### Study Design

We conducted a cross-sectional analysis by pooling five years of data (2017 to 2021) from the National Emergency Medical Services (EMS) Information System (NEMSIS). The NEMSIS is the national database of all EMS cases across U.S. States and territories [32]. Between 2017 and 2021, the number of states and territories that reported their EMS statistics to NEMSIS and permitted its use for research increased from 35 to 53 and the number of 9-1-1 events captured in the NEMSIS data increased from 7,907,829 to 48,982,990 [33].

### Inclusion and Exclusion Criteria

Between 2017 and 2021, 157,115,593 persons were managed following an EMS activation (Figure 1). We identified the older adult population (age 65 years and older) (n=58,272,048). We further restricted the population to age 65 years and older road users that sustained motor vehicle crash injuries using the International Classification of Disease version 10 (ICD-10) codes V00 to V79 (n= 489,565). We excluded cases whose substance use status was coded as “not applicable” (n=19,519; 4% of 489,565). Thereafter, we excluded cases whose injury status was not reported (n=213,897; 45.5% of 470,046). These unreported cases represent patients who either canceled the 9-1-1 call, refused care, or were evaluated but no treatment or transport was required. Also, we performed a listwise deletion for cases whose missingness was less than one percent (n=1,203; 0.5% of 256,149) and when the crash response time was greater than 60 minutes (n=1,103; 0.4% of 256,149). We excluded cases whose EMS response time exceeded 60 minutes, consistent with an earlier study [26]. These outlier cases are typically associated with unique environmental conditions such as tornadoes [34–36]. The final analytic data, therefore, was a total of 253,933 older adult road users who sustained motor vehicle injuries.

**Figure 1:**
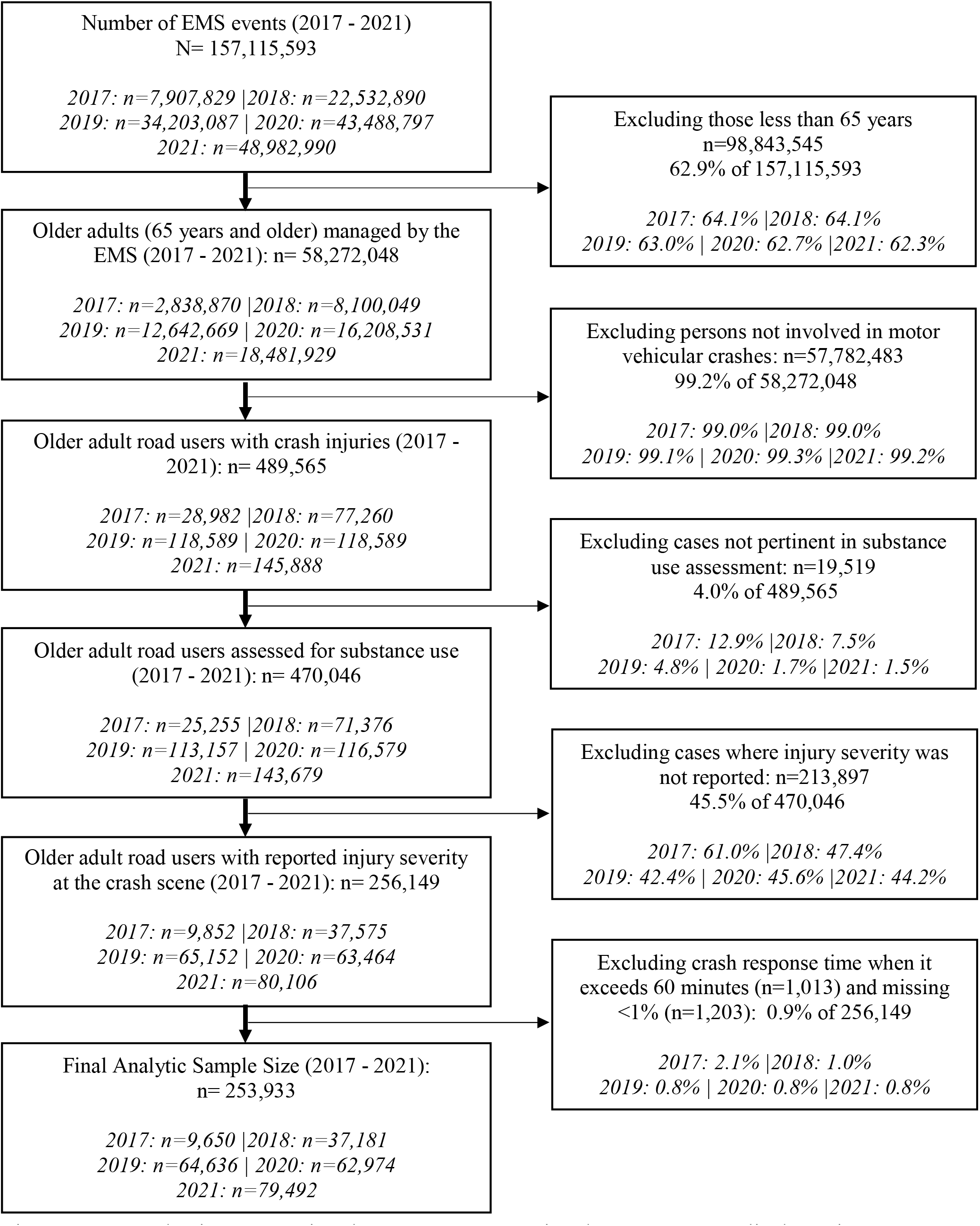
Data selection steps using the 2017 to 2021 National Emergency Medical Service (EMS) Information System database.

### Injury Severity

Our outcome measure, injury severity status, is a four-point categorical variable: low acuity, emergent, critical, and fatal injury [37]. Patients with low acuity injuries have injuries with a low probability of worsening or developing serious complications in the absence of intervention. Patients with emergent injuries have the potential of worsening if intervention is not initiated quickly. Patients with critical injuries have life-threatening injuries at high mortality risk if intervention is not initiated immediately.

Finally, those with fatal injuries either died at the crash scene or while in transit to the hospital. These categorizations were made by EMS providers at the crash scene using the Model of the Clinical Practice of Emergency Medicine [38,39].

### Substance Use

The main predictor variable is the presence or absence of substance use. We defined substance use using the variable eHistory.17, representing alcohol/drug use indicators [40]. The NEMSIS defines substance use in seven categories: 1) alcohol containers/paraphernalia at the scene, 2) drug paraphernalia at the scene, 3) patient admits to alcohol use, 4) patient admits to drug use, 5) positive level (of alcohol or drugs) known from law enforcement or hospital record, 6) smell of alcohol on breath, 7) none reported. We defined the presence of substance use at the time of the crash event as cases in the first to sixth categories while the absence of substance use at the time of the crash was defined with cases coded in the seventh category [40]. In the NEMSIS documentation, an individual can be assigned multiple categories. We, therefore, defined cases of “alcohol use only” using categories 1, 3, 5, and 6 and “drug use only” using categories 2 and 4.

### Patient and Injury Characteristics

We controlled for age, sex, race/ethnicity, road user type, anatomical injured region, rurality/urbanicity, environmental location of the crash, the time of the day of the crash, and the EMS response time. We selected these patient and injury characteristics a priori from the literature [14,24,26]. Age was measured as a three-level categorical variable while sex was measured as a binary variable. We defined race/ethnicity in four categories: non-Hispanic White, non-Hispanic Black, Hispanic, and other races. Road user type was measured in four categories using the ICD-10 codes: car occupants (V40 – V49), pedestrians (V00 – V09), two/three-wheel vehicle occupants (V10 – V39), and occupants of trucks, buses, industrial vehicles (V50 – V79). We defined the injured anatomic region in five categories: injury to the head and neck, abdomen and genitals, chest and back, upper and lower limbs (extremities), and multiple body injuries. NEMSIS reports the geographical location as a four-point categorical variable: wilderness, rural, suburban, and urban, using the United States Department of Agriculture Urban influence codes [37]. We recoded this variable into three categories: rural/wilderness (hereafter referred to as rural), suburban, and urban.

Following the recommended NEMSIS incident location categorization,[41] we defined the environmental location of the crash in three categories, using the ICD-10 codes: parking lots/sidewalks (Y92.48, including Y92.480 - Y.92.489), streets/highways/paved roads (Y92.41, including Y92.410 – Y92.419, and Y.92.4), and other locations. The other crash locations include places of residence, businesses, stores, recreational areas, schools, and places not otherwise specified. We defined the time of the crash injury using a proxy measure – the time the 9-1-1 call was initiated. The time of crash injury was defined in four categories: morning rush hour period, afternoon rush hour period, nighttime, and other hours. Using a recently published meta-analysis as a guide,[42] the morning and afternoon rush hour periods were defined as crash injuries sustained between 6 to 9 am and 3 to 7 pm, respectively. Nighttime crashes were defined as crash injuries occurring between 12 midnight and 5 am. We defined the EMS response time as the duration from initiation to arrival at the crash scene and it was measured as a four-point categorical variable: less than nine minutes, nine to 17.59 minutes, 18 to 26.59 minutes, and 27 minutes or higher.

### Handling of Missing Data

We encountered missing values in the following variables: substance use (18.0%), race/ethnicity (30.5%), environmental location of crash (1.6%), and anatomical injured region (25.7%). We performed multiple imputations for missing data, using the multiple imputations with chained equation (MICE) after justifying that missingness was at random [43]. Additionally, NEMSIS had advised researchers not to assume that missingness in the NEMSIS data is “Not Missing at Random”,[44] further stressing the need to perform some measures of missing data analysis whenever such missingness is encountered. The MICE model was strengthened using injury severity, age, sex, crash response time, road user type, and the time of the crash as predictors. We performed 100 iterations, generated 100 predicted values for all missing values, and assigned the final value using the mean of the predicted values, consistent with earlier literature on multiple imputations [45,46].

### Analysis

We computed the frequency distribution of demographic, injury, crash, and substance use characteristics. We assessed differences across injury severity status and rurality/urbanicity using chi-square statistics. We performed a partially proportional ordinal logistic regression[47] to assess the odds of worse injury outcomes – (critical, emergent, and fatal injuries) and computed the predicted probabilities of substance use-associated injury severity. The decision to use a partially proportional ordinal logistic regression, as opposed to a proportionally ordinal regression was based on the violation of the parallel lines assumption evidenced by a significant Brant test [47]. Also, we performed the interaction analysis between substance use and rurality/urbanicity and we reported the predicted probabilities of each substance use-related injury severity category in rural, suburban, and urban areas. Data were analyzed using SAS 9.4[48] and STATA version 17 [49].

## Results

A total of 253,933 older adults met our inclusion and exclusion criteria (Table 1). The majority of the population was between 65 and 74 years (62%), female (51%), non-Hispanic Whites (72%), and car occupants (76%). Thirty-six percent of the sample population sustained injuries to the chest and back. The crash injuries occurred mostly in urban areas (83%) with 6% of the injuries occurring at parking lots/sidewalks, and 16% occurring during the afternoon rush hour period. Approximately 75% of the older adults experienced an EMS response time of less than nine minutes. Substance use was identified in approximately 3% of the sample population with cases of only alcohol or only drug impairments being 2.9% and 0.4%, respectively. Furthermore, 67% of the sample population had low acuity injuries, 25%, and 6% sustained emergent and critical injuries and 1% died. Age, sex, race/ethnicity, road user type, the anatomical injured region, geographical location, environmental location of the crash, time of the day, EMS response time, and measures of substance use were significantly associated with pre-hospital crash injury severity(p<0.001).

**Table 1:** Frequency distribution and summary statistics of the demographic, crash, injury, and substance use characteristics of the study population (N=253,933)

| Variables | Frequency (%)<br>(N=253,933) | Low Acuity<br>n=170,805<br>(67.3%) | Emergent<br>n=64,226<br>(25.3%) | Critical<br>n=16,070<br>(6.3%) | Fatal<br>n=2,832<br>(1.1%) | p-value* |
| --- | --- | --- | --- | --- | --- | --- |
| Age Categories |  |  |  |  |  |  |
| 65 – 74 years | 157,496 (62.0) | 106,854 (62.6) | 39,336 (61.3) | 9,602 (59.8) | 1,704 (60.2) | <0.001 |
| 75 – 84 years | 72,969 (28.7) | 49,012 (28.7) | 18,685 (29.1) | 4,475 (27.9) | 797 (28.1) |  |
| 85 years and older | 23,468 (9.2) | 14,939 (8.7) | 6,205 (9.7) | 1,993 (12.4) | 331 (11.7) |  |
| Sex |  |  |  |  |  |  |
| Male | 124,829 (49.2) | 77,801 (45.6) | 34,911 (54.4) | 10,207 (63.5) | 1,910 (67.4) | <0.001 |
| Female | 129,104 (50.8) | 93,004 (54.4) | 29,315 (45.6) | 5,863 (36.5) | 922 (32.6) |  |
| Race/Ethnicity |  |  |  |  |  |  |
| Non-Hispanic White | 181,463 (71.5) | 116,325 (68.1) | 49,763 (77.5) | 13,033 (81.1) | 2,342 (82.7) | <0.001 |
| Non-Hispanic Black | 54,008 (21.3) | 41,292 (24.2) | 10,363 (16.1) | 2,018 (12.6) | 335 (11.8) |  |
| Hispanic | 13,009 (5.1) | 9,328 (5.5) | 2,837 (4.4) | 714 (4.4) | 130 (4.6) |  |
| Other Races | 5,453 (2.1) | 3,860 (2.3) | 1,263 (2.0) | 305 (1.9) | 25 (0.9) |  |
| Road User Type |  |  |  |  |  |  |
| Car Occupant | 192,450 (75.8) | 136,636 (80.0) | 44,815 (69.8) | 9,319 (58.0) | 1,680 (59.3) | <0.001 |
| Pedestrian | 24,900 (9.8) | 13,848 (8.1) | 7,651 (11.9) | 2,902 (18.0) | 499 (17.7) |  |
| In Two/Three Wheel Vehicles | 25,246 (9.9) | 13,548 (7.9) | 8,590 (13.4) | 2,779 (17.3) | 329 (11.6) |  |
| In Truck/Buses/Industrial Vehicle | 11,337 (4.5) | 6,773 (10.0) | 3,170 (4.9) | 1,070 (6.7) | 324 (11.4) |  |
| Anatomical Injured Region |  |  |  |  |  |  |
| Head and Neck | 51,940 (20.4) | 29,971 (17.6) | 16,300 (25.4) | 5,024 (31.3) | 645 (22.8) | <0.001 |
| Chest and Back | 91,892 (36.2) | 69,910 (40.9) | 19,353 (30.1) | 2,517 (15.7) | 112 (4.0) |  |
| Abdomen and Genitals | 4,585 (1.8) | 2,775 (1.6) | 1,407 (2.2) | 392 (2.4) | 11 (0.4) |  |
| Extremities | 41,537 (16.4) | 29,966 (17.5) | 9,912 (15.4) | 1,621 (10.1) | 38 (1.3) |  |
| Multiple Body Injuries | 63,979 (25.2) | 38,183 (22.4) | 17,254 (26.9) | 6,516 (40.5) | 2,026 (71.5) |  |
| Geographical Location |  |  |  |  |  |  |
| Urban | 209,791 (82.6) | 144,539 (84.6) | 51,184 (79.7) | 12,107 (75.3) | 1,961 (69.2) | <0.001 |
| Suburban | 20,294 (8.0) | 12,601 (7.4) | 5,403 (8.4) | 1,875 (11.7) | 415 (14.7) |  |
| Rural | 23,848 (9.4) | 13,665 (8.0) | 7,639 (11.9) | 2,088 (13.0) | 456 (16.1) |  |
| Environmental Location of Crash |  |  |  |  |  |  |
| Street or Highways | 187,692 (73.9) | 128,242 (75.1) | 45,892 (71.5) | 11,159 (69.5) | 2,399 (84.7) | <0.001 |
| Parking lots/Sidewalks | 15,472 (6.1) | 10,687 (6.3) | 3,729 (5.8) | 921 (5.7) | 135 (4.8) |  |
| Other locations | 50,769 (20.0) | 31,876 (18.6) | 14,605 (22.7) | 3,990 (24.8) | 298 (10.5) |  |
| Time of the Day |  |  |  |  |  |  |
| Morning Rush Hour | 16,683 (6.6) | 11,472 (6.7) | 3,969 (6.2) | 1,030 (6.4) | 212 (7.5) | <0.001 |
| Afternoon Rush Hour | 40,766 (16.1) | 27,529 (16.1) | 10,136 (15.8) | 2,691 (16.8) | 410 (14.5) |  |
| Nighttime Driving | 11,138 (4.4) | 7,228 (4.2) | 2,926 (4.6) | 797 (5.0) | 187 (6.6) |  |
| Other Hours | 185,346 (73.0) | 124,576 (72.9) | 47,195 (73.5) | 11,552 (71.9) | 2,023 (71.4) |  |
| EMS Response Time |  |  |  |  |  |  |
| 0 – 8.59 minutes | 190,619 (75.1) | 130,122 (76.2) | 47,338 (73.7) | 11,213 (69.8) | 1,946 (68.7) |  |
| 9 – 17.59 minutes | 50,502 (19.9) | 33,264 (19.5) | 12,990 (20.2) | 3,524 (21.9) | 724 (25.6) |  |
| 18 – 26.59 minutes | 9,068 (3.6) | 5,377 (3.1) | 2,660 (4.2) | 908 (5.7) | 123 (4.3) |  |
| 27 minutes or higher | 3,744 (1.5) | 2,042 (1.2) | 1,238 (1.9) | 425 (2.6) | 39 (1.4) |  |
| Substance Use |  |  |  |  |  |  |
| Yes | 7,948 (3.1) | 4,337 (2.5) | 2,658 (4.1) | 883 (5.5) | 70 (2.5) | <0.001 |
| No | 245,985 (96.9) | 166,468 (97.5) | 61,568 (95.9) | 15,187 (94.5) | 2,762 (97.5) |  |
| Alcohol Use Alone |  |  |  |  |  |  |
| Yes | 7,239 (2.9) | 3,974 (2.3) | 2,419 (3.8) | 786 (4.9) | 60 (2.1) | <0.001 |
| No | 246,694 (97.1) | 166,831 (97.7) | 61,807 (96.2) | 15,284 (95.1) | 2,772 (97.9) |  |
| Drug Use Alone |  |  |  |  |  |  |
| Yes | 970 (0.4) | 596 (0.4) | 273 (0.4) | 91 (0.6) | 10 (0.4) | <0.001 |
| No | 252,963 (99.6) | 170,209 (99.6) | 63,953 (99.6) | 15,979 (99.4) | 2,822 (99.6) |  |

\*Statistical differences in low acuity, emergent, and critical categories across all variables except Revised Trauma Score were
assessed using chi-square; \*\*Mean differences in Revised Trauma Score were assessed using the one-way ANOVA, and median
differences were assessed using the Kruskal-Wallis test

Between 2017 and 2021, the proportion of emergent injuries ranged from 25.0 - 27.3%, while the proportion of critical and fatal injuries ranged between 5.7% - 8.4%, and 1.0 - 1.2%, respectively (Figure 2). Among older adults with emergent injuries, the proportions in urban areas ranged between 24.0 - 26.3%, while the proportions in suburban and rural areas ranged between 25.6 - 30.9%, and 31.4 - 33.5%, respectively (p<0.001). Also, among older adults with critical injuries, the proportions ranged between 5.2 - 7.8% in urban areas, and in suburban and rural areas, the proportions ranged between 8.6 - 13.2% and 8.0 - 9.4%, respectively (p<0.001). Furthermore, among older adults with fatal injuries, the proportions in urban areas ranged between 0.8 - 1.1%, and in suburban and rural areas, the proportions ranged between 1.8 - 2.3%, and 1.5 - 2.2%, respectively (p<0.001). While there was a decline in emergent and critical injuries between 2017 and 2018, the proportions of emergent, critical, and fatal injuries gradually increased from 2019 to 2021 in rural, suburban, and urban areas (p<0.001).

**Figure 2:**
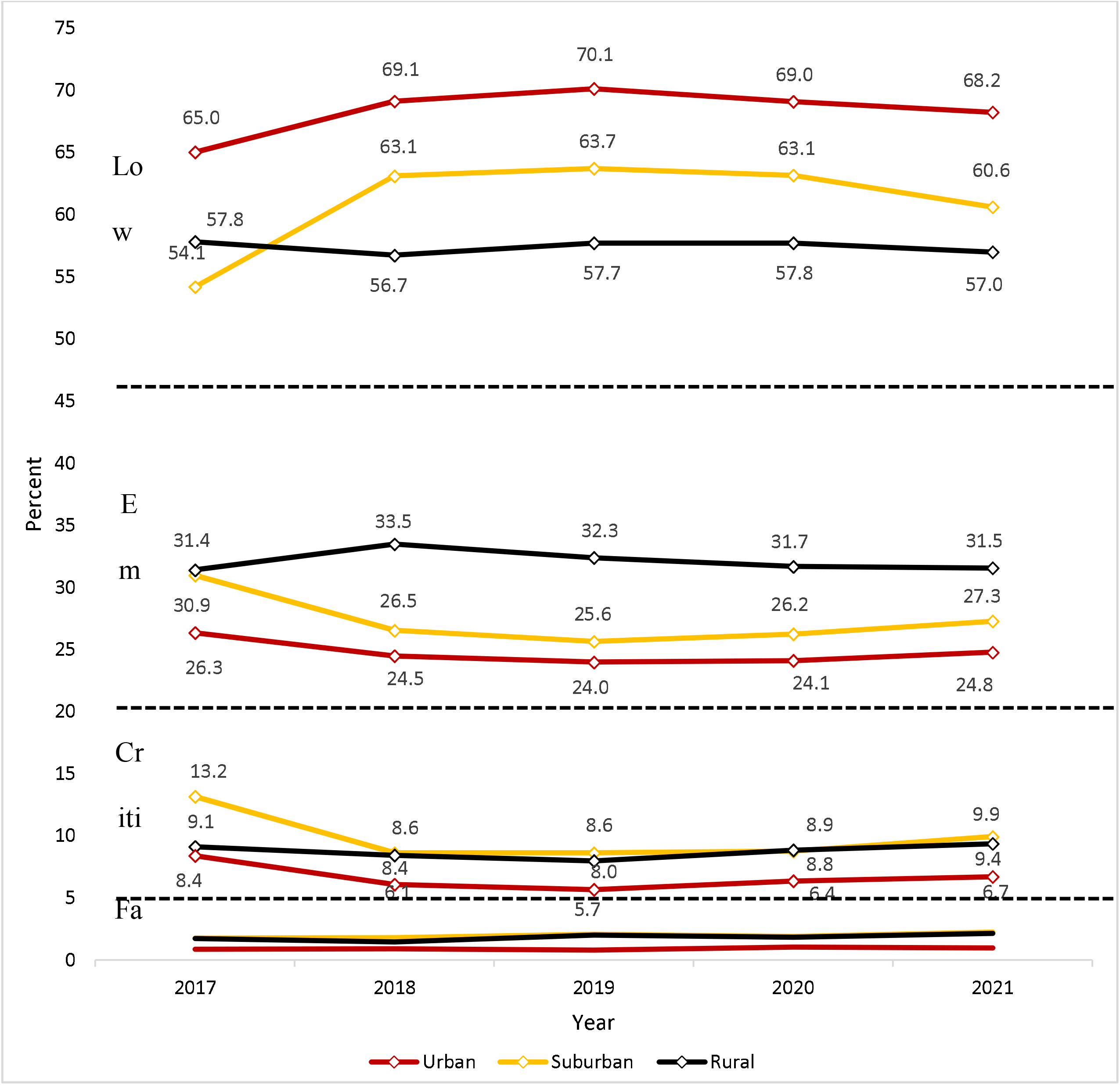
Trend of the proportion of low acuity, emergent, critical, and fatal crash injuries among older adult road users across rural, suburban, and urban areas between 2017 and 2021

There were significant differences in the age and sex of older adults who sustained crash injuries in rural, suburban, and urban areas (Table 2). The proportion of non-Hispanic Whites who sustained crash injuries in urban areas was 68% while the proportions in suburban and rural areas were 86% and 87%, respectively (p<0.001). Also, the proportion of occupants of buses and trucks involved in crash injuries was 4% in urban areas, and in suburban and rural areas, the proportions were 8% and 9%, respectively. The proportion of older adults who sustained multiple body injuries was 25% in urban areas, and in suburban and rural areas, each proportion was 28% (p<0.001). The proportion of older adults who sustained injuries at parking lots and sidewalks was 7% in urban areas, and in each suburban and rural area, the proportion was 4% (p<0.001). The proportion of older adults who experienced EMS response time of less than nine minutes was 77% in urban areas, and in suburban and rural areas, the proportions were 67% and 63%, respectively (p<0.001). Although the proportion of drug use in urban areas was significantly higher compared to suburban and rural areas (p<0.001), there were no differences in alcohol use in urban, suburban, and rural areas.

**Table 2:**
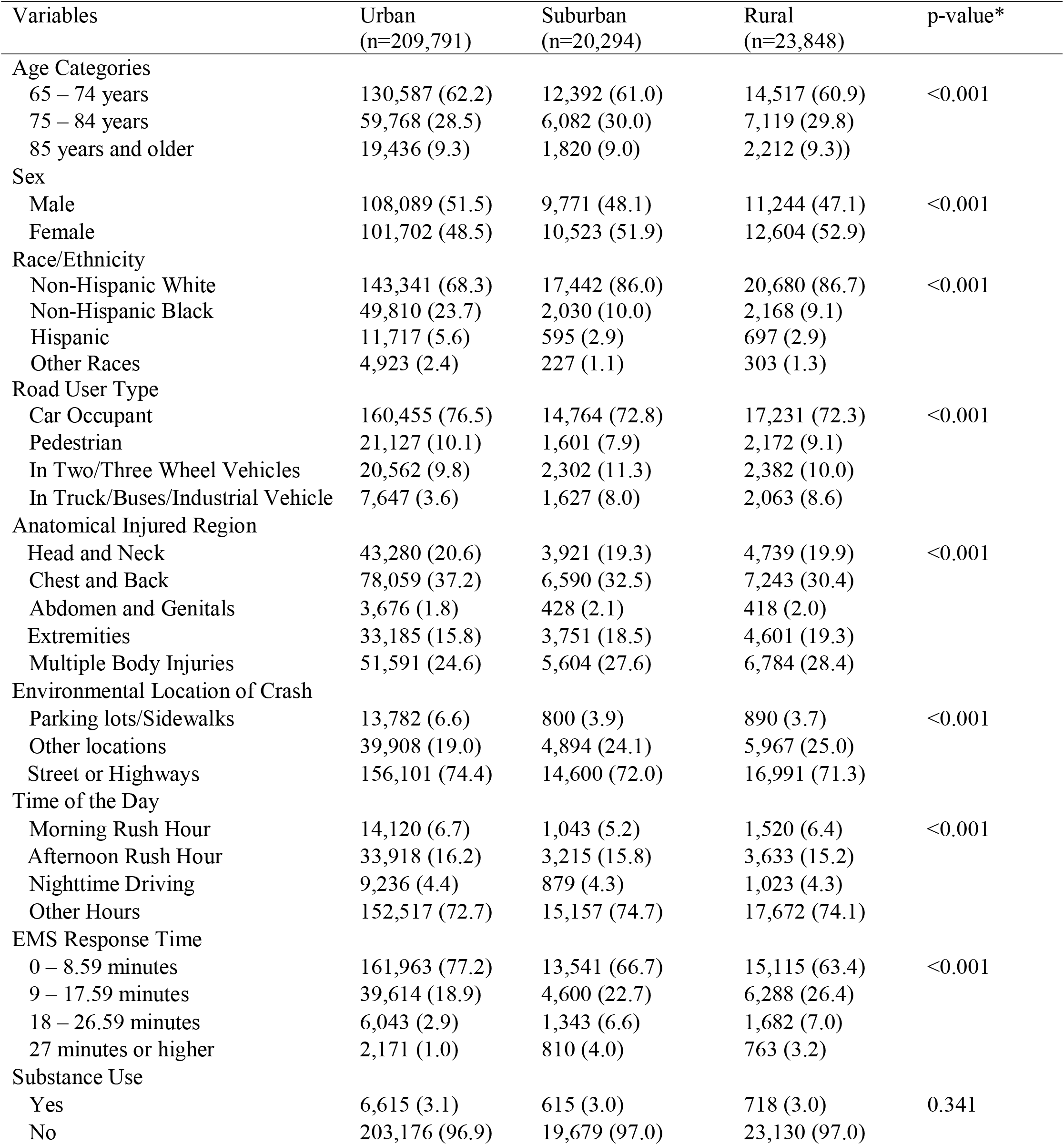
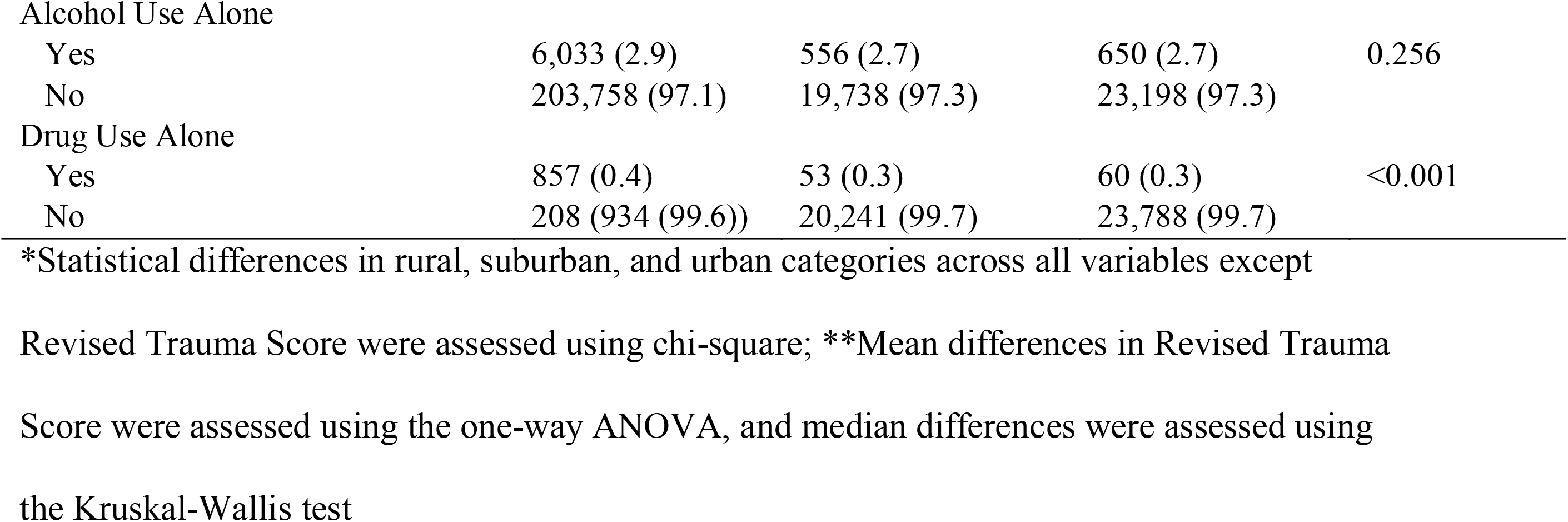
Rural-urban differences in the frequency distribution and summary statistics of the demographic, crash, injury and substance use characteristics of the study population (N=253,933)

| Variables | Urban<br>(n=209,791) | Suburban<br>(n=20,294) | Rural<br>(n=23,848) | p-value* |
| --- | --- | --- | --- | --- |
| Age Categories |  |  |  |  |
| 65 – 74 years | 130,587 (62.2) | 12,392 (61.0) | 14,517 (60.9) | <0.001 |
| 75 – 84 years | 59,768 (28.5) | 6,082 (30.0) | 7,119 (29.8) |  |
| 85 years and older | 19,436 (9.3) | 1,820 (9.0) | 2,212 (9.3)) |  |
| Sex |  |  |  |  |
| Male | 108,089 (51.5) | 9,771 (48.1) | 11,244 (47.1) | <0.001 |
| Female | 101,702 (48.5) | 10,523 (51.9) | 12,604 (52.9) |  |
| Race/Ethnicity |  |  |  |  |
| Non-Hispanic White | 143,341 (68.3) | 17,442 (86.0) | 20,680 (86.7) | <0.001 |
| Non-Hispanic Black | 49,810 (23.7) | 2,030 (10.0) | 2,168 (9.1) |  |
| Hispanic | 11,717 (5.6) | 595 (2.9) | 697 (2.9) |  |
| Other Races | 4,923 (2.4) | 227 (1.1) | 303 (1.3) |  |
| Road User Type |  |  |  |  |
| Car Occupant | 160,455 (76.5) | 14,764 (72.8) | 17,231 (72.3) | <0.001 |
| Pedestrian | 21,127 (10.1) | 1,601 (7.9) | 2,172 (9.1) |  |
| In Two/Three Wheel Vehicles | 20,562 (9.8) | 2,302 (11.3) | 2,382 (10.0) |  |
| In Truck/Buses/Industrial Vehicle | 7,647 (3.6) | 1,627 (8.0) | 2,063 (8.6) |  |
| Anatomical Injured Region |  |  |  |  |
| Head and Neck | 43,280 (20.6) | 3,921 (19.3) | 4,739 (19.9) | <0.001 |
| Chest and Back | 78,059 (37.2) | 6,590 (32.5) | 7,243 (30.4) |  |
| Abdomen and Genitals | 3,676 (1.8) | 428 (2.1) | 418 (2.0) |  |
| Extremities | 33,185 (15.8) | 3,751 (18.5) | 4,601 (19.3) |  |
| Multiple Body Injuries | 51,591 (24.6) | 5,604 (27.6) | 6,784 (28.4) |  |
| Environmental Location of Crash |  |  |  |  |
| Parking lots/Sidewalks | 13,782 (6.6) | 800 (3.9) | 890 (3.7) | <0.001 |
| Other locations | 39,908 (19.0) | 4,894 (24.1) | 5,967 (25.0) |  |
| Street or Highways | 156,101 (74.4) | 14,600 (72.0) | 16,991 (71.3) |  |
| Time of the Day |  |  |  |  |
| Morning Rush Hour | 14,120 (6.7) | 1,043 (5.2) | 1,520 (6.4) | <0.001 |
| Afternoon Rush Hour | 33,918 (16.2) | 3,215 (15.8) | 3,633 (15.2) |  |
| Nighttime Driving | 9,236 (4.4) | 879 (4.3) | 1,023 (4.3) |  |
| Other Hours | 152,517 (72.7) | 15,157 (74.7) | 17,672 (74.1) |  |
| EMS Response Time |  |  |  |  |
| 0 – 8.59 minutes | 161,963 (77.2) | 13,541 (66.7) | 15,115 (63.4) | <0.001 |
| 9 – 17.59 minutes | 39,614 (18.9) | 4,600 (22.7) | 6,288 (26.4) |  |
| 18 – 26.59 minutes | 6,043 (2.9) | 1,343 (6.6) | 1,682 (7.0) |  |
| 27 minutes or higher | 2,171 (1.0) | 810 (4.0) | 763 (3.2) |  |
| Substance Use |  |  |  |  |
| Yes | 6,615 (3.1) | 615 (3.0) | 718 (3.0) | 0.341 |
| No | 203,176 (96.9) | 19,679 (97.0) | 23,130 (97.0) |  |
| Alcohol Use Alone |  |  |  |  |
| Yes | 6,033 (2.9) | 556 (2.7) | 650 (2.7) | 0.256 |
| No | 203,758 (97.1) | 19,738 (97.3) | 23,198 (97.3) |  |
| Drug Use Alone |  |  |  |  |
| Yes | 857 (0.4) | 53 (0.3) | 60 (0.3) | <0.001 |
| No | 208 (934 (99.6)) | 20,241 (99.7) | 23,788 (99.7) |  |

\*Statistical differences in rural, suburban, and urban categories across all variables except
Revised Trauma Score were assessed using chi-square; \*\*Mean differences in Revised Trauma Score were assessed using the one-way ANOVA, and median differences were assessed using the Kruskal-Wallis test

Compared to low acuity injury, the unadjusted odds of worse injury severity were higher with increasing age, among males, pedestrians, occupants of two- or three-wheeled vehicles, and occupants of buses and trucks (Table 3). Worse injury severity was also higher among those with injuries to the head and neck, those that were involved in nighttime driving, and with increasing EMS crash response time. Compared to injuries in urban areas, injuries that occurred in rural (OR: 1.65; 95% CI: 1.61 - 1.70) and suburban areas (OR: 1.35; 95% CI: 1.31 - 1.39) were associated with increased odds of worse injury severity. Substance use was associated with 1.7 (95% CI: 1.67 - 1.82) times the odds of worse injury severity. After adjusting for potential confounders, substance use was associated with 1.35 times the adjusted odds of worse injury severity (95% CI: 1.29 - 1.42). Alcohol use alone was associated with 1.32 (95% CI: 1.26 - 1.39) times the adjusted odds of worse injury severity while drug use alone was associated with 1.15 (95% CI: 1.01 - 1.32) times the adjusted odds of worse injury severity.

**Table 3:** Unadjusted and adjusted odds ratio of worse injury severity (critical, emergent, death vs. low acuity) associated with the demographic, crash, injury, and substance use characteristics among older adults

| Variables | Emergent/Critical/Fatal vs. Low Acuity |  |  |  |
| --- | --- | --- | --- | --- |
|  | Univariate Model | Substance Use Model | Alcohol Alone Model | Drugs Alone Model |
|  | Unadjusted Odds Ratio (95% CI) | Adjusted Odds Ratio (95% CI)*** | Adjusted Odds Ratio (95% CI)*** | Adjusted Odds Ratio (95% CI)*** |
| Substance Use |  |  |  |  |
| Yes | **1.74 (1.67 – 1.82)** | **1.35 (1.29 – 1.42)** |  |  |
| No | Ref | Ref |  |  |
| Alcohol Use Alone |  |  |  |  |
| Yes | **1.72 (1.64 – 1.80)** |  | **1.32 (1.26 – 1.39)** |  |
| No | Ref |  | Ref |  |
| Drug Use Alone |  |  |  |  |
| Yes | **1.29 (1.13 – 1.47)** |  |  | **1.15 (1.01 – 1.32)** |
| No | Ref |  |  | Ref |
| Age Categories |  |  |  |  |
| 85 years and older | **1.20 (1.17 – 1.24)** | **1.18 (1.15 – 1.22)** | **1.18 (1.15 – 1.22)** | **1.18 (1.15 – 1.22)** |
| 75 – 84 years | **1.03 (1.01 – 1.05)** | **1.04 (1.02 – 1.06)** | **1.04 (1.02 – 1.06)** | **1.04 (1.02 – 1.06)** |
| 65 – 74 years | Ref | Ref | Ref | Ref |
| Sex |  |  |  |  |
| Male | **1.56 (1.53 – 1.58)** | **1.29 (1.27 – 1.32)** | **1.30 (1.27 – 1.32)** | **1.31 (1.28 – 1.33)** |
| Female | Ref | Ref | Ref | Ref |
| Race/Ethnicity |  |  |  |  |
| Non-Hispanic Black | **0.55 (0.54 – 0.56)** | **0.60 (0.59 – 0.62)** | **0.60 (0.59 – 0.62)** | **0.60 (0.59 – 0.62)** |
| Hispanic | **0.70 (0.68 – 0.73)** | **0.72 (0.69 – 0.75)** | **0.72 (0.69 – 0.75)** | **0.72 (0.69 – 0.75)** |
| Other Races | **0.74 (0.69 – 0.78)** | **0.77 (0.73 – 0.82)** | **0.77 (0.73 – 0.82)** | **0.77 (0.72 – 0.82)** |
| Non-Hispanic White | Ref | Ref | Ref | Ref |
| Road User Type |  |  |  |  |
| Pedestrian | **1.95 (1.90 – 2.01)** | **1.95 (1.90 – 2.02)** | **1.96 (1.90 – 2.02)** | **1.97 (1.91 – 2.03)** |
| Two/Three Wheels | **2.11 (2.06 – 2.17)** | **1.93 (1.88 – 1.99)** | **1.93 (1.88 – 1.99)** | **1.94 (1.88 – 1.99)** |
| Truck/Buses/Heavy | **1.64 (1.59 – 1.71)** | **1.47 (1.44 – 1.53)** | **1.47 (1.44 – 1.53)** | **1.48 (1.42 – 1.53)** |
| Vehicle |  |  |  |  |
| Cars | Ref | Ref | Ref | Ref |
| Anatomical Injured Region |  |  |  |  |
| Head and Neck | **1.08 (1.06 – 1.11)** | **1.13 (1.10 – 1.15)** | **1.13 (1.10 – 1.15)** | **1.13 (1.10 – 1.15)** |
| Chest and Back | **0.47 (0.46 – 0.48)** | **0.48 (0.47 – 0.50)** | **0.48 (0.47 – 0.50)** | **0.48 (0.47 – 0.49)** |
| Abdomen and Genitals | 0.97 (0.91 – 1.03) | 1.01 (0.95 – 1.08) | 1.01 (0.95 – 1.08) | 1.01 (0.94 – 1.07) |
| Extremities | **0.57 (0.56 – 0.59)** | **0.50 (0.49 – 0.52)** | **0.50 (0.49 – 0.52)** | **0.50 (0.49 – 0.52)** |
| General Body | Ref | Ref | Ref | Ref |
| Geographical Location |  |  |  |  |
| Rural | **1.65 (1.61 – 1.70)** | **1.44 (1.40 – 1.48)** | **1.44 (1.40 – 1.48)** | **1.44 (1.40 – 1.48)** |
| Suburban | **1.35 (1.31 – 1.39)** | **1.19 (1.16 – 1.23)** | **1.19 (1.16 – 1.23)** | **1.19 (1.16 – 1.23)** |
| Urban | Ref | Ref | Ref | Ref |
| Environmental Location |  |  |  |  |
| Parking lots/Sidewalks | 0.97 (0.93 – 1.00) | **0.88 (0.85 – 0.91)** | **0.88 (0.85 – 0.91)** | **0.88 (0.84 – 0.91)** |
| Other locations | **1.28 (1.25 – 1.30)** | **1.05 (1.03 – 1.08)** | **1.05 (1.03 – 1.08)** | **1.05 (1.03 – 1.08)** |
| Street or Highways | Ref | Ref | Ref | Ref |
| Time of the Day |  |  |  |  |
| Morning Rush Hour | **0.93 (0.90 – 0.96)** | **0.92 (0.89 – 0.95)** | **0.92 (0.89 – 0.95)** | **0.91 (0.88 – 0.95)** |
| Afternoon Rush Hour | 0.99 (0.96 – 1.01) | **0.97 (0.95 – 0.99)** | **0.98 (0.95 – 0.99)** | **0.98 (0.95 – 0.99)** |
| Nighttime Driving | **1.11 (1.07 – 1.15)** | **1.07 (1.02 – 1.11)** | **1.07 (1.02 – 1.11)** | **1.07 (1.03 – 1.12)** |
| Other Hours | Ref | Ref | Ref | Ref |
| EMS Response Time |  |  |  |  |
| 27 minutes or higher | **1.79 (1.68 – 1.91)** | **1.44 (1.34 – 1.54)** | **1.44 (1.34 – 1.54)** | **1.43 (1.34 – 1.54)** |
| 18 – 26.59 minutes | **1.48 (1.41 – 1.54)** | **1.30 (1.24 – 1.36)** | **1.30 (1.24 – 1.36)** | **1.30 (1.24 – 1.36)** |
| 9 – 17.59 minutes | **1.11 (1.09 – 1.14)** | **1.07 (1.05 – 1.09)** | **1.07 (1.05 – 1.09)** | **1.07 (1.05 – 1.09)** |
| 0 – 8.59 minutes | Ref | Ref | Ref | Ref |

\*Univariate logistic regression was used to perform the analysis; \*\*Partially proportional ordinal
logistic regression was used to perform the analysis; \*\*\*Adjusted model consists of all listed variables.

The predicted probability of substance use-associated emergent injury was 32.0% (95% CI: 30.9 - 33.0) and the predicted probability increased step-wisely from urban (31.2%; 95% CI: 30.2 - 32.3) to suburban (33.5; 95% CI: 29.8 - 37.1) and rural areas (36.5%; 95% CI: 33.0 - 39.9) (p<0.001) (Figure 3). Also, the predicted probability of substance use-associated critical injury was 6.0% (95% CI: 5.6 - 6.4). The predicted probability was lowest in urban areas (5.7%; 95% CI: 5.3 - 6.2) and in suburban and rural areas, the values were 8.3% (95% CI: 6.7 - 10.0) and 7.1% (95% CI: 5.7 - 8.5), respectively (p<0.001). Furthermore, the predicted probability of substance use-associated fatal injury was 0.2% (95% CI: 0.1 - 0.2). The predicted probability was 0.2% (95% CI: 0.1 - 0.2) in urban areas, and in suburban and rural areas, the values were 0.5% (95% CI: 0.2 - 0.7) and 0.4% (95% CI: 0.2 - 0.6), respectively (p<0.001).

**Figure 3:**
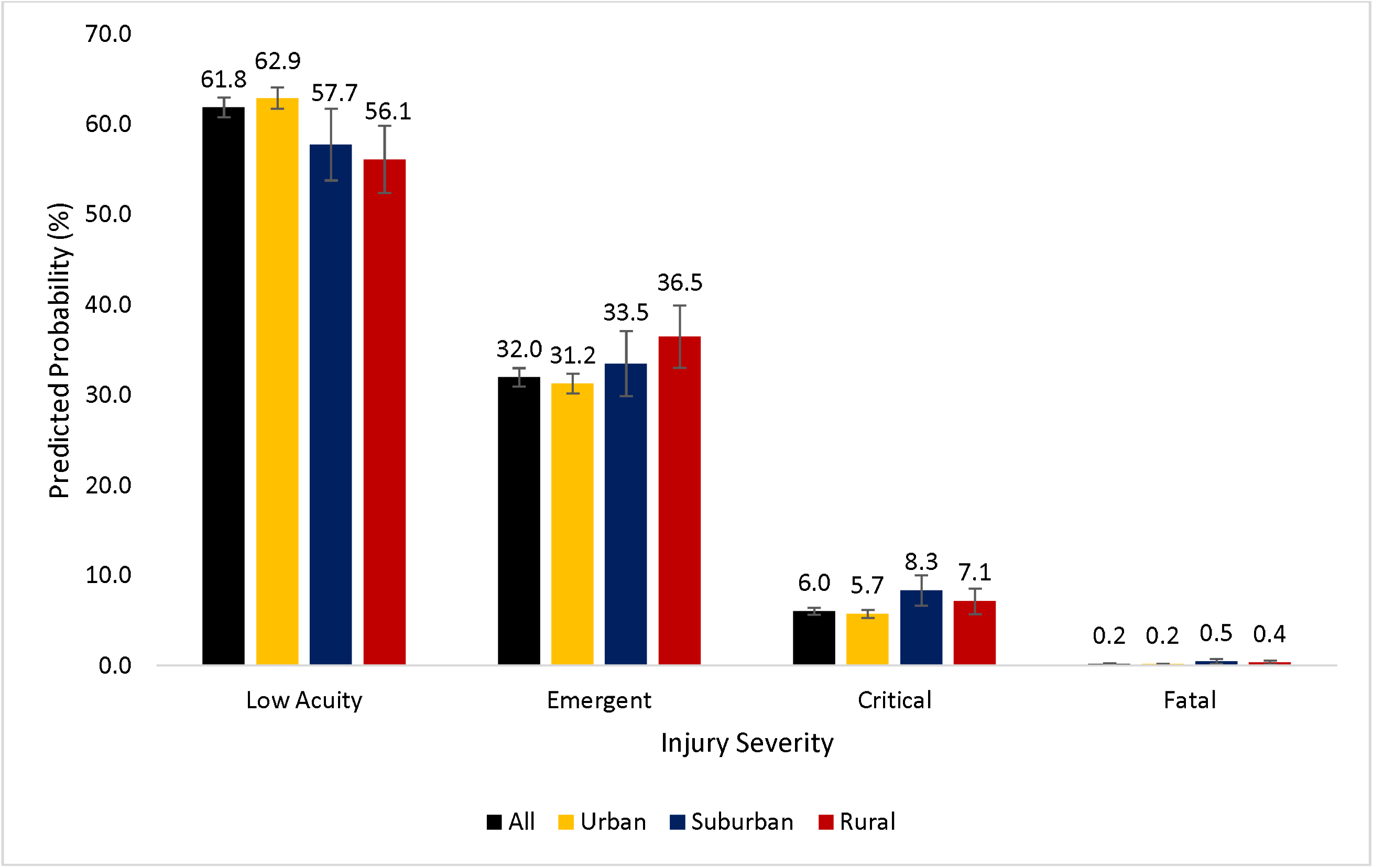
Predicted probabilities of low acuity, emergent, critical, and fatal crash injuries among older adult road users in all areas, and in rural, suburban, and urban areas.

## Discussion

We present one of the few studies that demonstrate an association between substance use and crash injury severity among older adult road users. While earlier studies had reported the increased odds of fatal crash injuries from substance use,[13,14] our study provides additional context on the injury, crash response, and environmental predictors of worse injury severity among older adult road users. While substance use did not differ significantly between rural and urban settings, the likelihood of emergent, critical, and fatal injuries was significantly higher in rural/suburban areas compared to urban settings. Additionally, we report that four out of ten older road users involved in crash events who use alcohol or drugs are more likely to have emergent, critical, or fatal injuries and this likelihood is disproportionately higher in suburban and rural areas. Furthermore, an increasing trend in critical and fatal injury proportions between 2019 and 2021 in urban, suburban, and rural areas presents an area for public health intervention.

Earlier studies have reported the harmful effect of alcohol and drug use among older adults some of which include increased risk of falls,[50–52] cognitive impairment,[53,54] increased risk of alcohol or drug dependence, and worsening health conditions. Our report of the increased odds of worse injury severity from substance use adds to the extant literature on the harms associated with alcohol and/or drug use. The worse injury outcomes associated with substance use among older adult road users may be explained by the reduced efficiency in metabolizing alcohol and drugs, longer toxic exposure, and an attenuation of the injury response mechanism when older adults are exposed to acute and/or chronic use of alcohol or drugs [55]. Acute and chronic alcohol use is associated with orthostatic hypotension and hypertension,[56–58] respectively, and drugs such as opioids and benzodiazepines are central nervous system depressants [59,60]. Alcohol and drugs may further impair age-related physiologic response to acute trauma, hence increasing injury morbidity among older adults.

Earlier studies have reported no rural-urban differences in alcohol and drug consumption [61,62]. While we did not detect a difference in substance use between rural, suburban, and urban areas, the association between substance use and crash injury severity differed significantly. Also, while the predicted probabilities of low acuity injuries decreased from urban to suburban and rural areas, the predicted probabilities of emergent injuries had a step-wise increase from urban to suburban and rural. Additionally, suburban and rural areas had higher critical and fatal injury probabilities compared to urban areas. These observed variations may reflect the rural-urban differences in driving behavior and access to timely and appropriate emergency care. Earlier studies have reported increased speeding behavior among road users in rural areas[63] and non-use of seat belts among older adult drivers [64]. Rural areas also experience significantly prolonged response times,[24,26] increased deaths at the crash scene,[26] and increased proportions of crash fatalities [65–67].

Crash injuries among older adult road users are preventable and interventions aimed at reducing substance use among road users represent a strategy for reducing crash-related morbidity and mortality. Excluding crash events in 2017 and 2018, our report showed an increasing trend in critical and fatal injuries among older adult road users in rural, suburban, and urban areas. There is therefore a need for intensified and focused public health intervention in reducing older adult crash injury rates across the U.S. Preventing the interactions of several co-existing risk factors of crash occurrence is a strategy recommended by the Governors Highway Safety Administration [63]. For example, our study showed that night driving is associated with worse injury severity among older adult road users. However, intentionally increasing nighttime police presence and nighttime enforcement of speed limits on road sections and highways associated with high clusters of crash occurrence may further reduce crash occurrence at night. Preventing the interaction effect of risk factors requires improved data collection and the creation of more sensitive spatial and non-spatial models. Additionally, there is a need to extend substance use educational intervention to non-conventional research settings where older adults commonly gather. Such locations include medical clinics, senior centers, retirement communities, places of worship, and parks and recreation centers. Achieving behavioral change in alcohol and illicit drug use among older adults requires identifying motivators for change, using positive messaging techniques, encouraging peer support, and exercising patience [68,69]. Furthermore, primary care providers should educate older adults on the harm of driving when given prescription drugs.

This study has its limitations. As a cross-sectional design, causal inferences cannot be made. We were unable to control for other risky driving behaviors such as the non-use of seatbelts, distracted driving, and speeding because these variables were not captured in the NEMSIS. We did not adjust for the level of certification of crash scene EMS staff since this information is not part of the publicly-released data from the NEMSIS. Substance use, a heterogeneous term for alcohol, marijuana, narcotics, stimulants, depressants, benzodiazepines, and other illicit drugs is inconsistently screened across the U.S. Substance use screening differs across states with substantial under-reporting of drug use while driving,[70] falsely lowering the effect size we report. The gold standard for a diagnosis of substance use remains a serological assessment [71]. However, with some cases of substance use identified by self-reported measures and the presence of paraphernalia of alcohol and drugs, the possibility of misclassification bias is likely. Misclassification of the outcome measure is less likely since injury severity classification was based on a complex matrix and applied by a trained EMS staff. Despite these limitations, this study has several strengths, which include the generalizability of this study to older adults across the U.S. Also, this is one of the few studies that assess the rural-urban association of substance use and crash injury severity among older adult road users. The awareness of the association between substance use and crash injury severity as well as the rural-urban differences will improve interventions aimed at reducing crash involvement of older adult road users.

## Conclusion

Substance use is associated with worse crash injury severity among older adult road users. Despite no significant difference in rural-urban proportions of substance use among older adults, emergent injuries increase from urban to rural areas. Increasing trends in critical and fatal older adult injuries should motivate urgent public health interventions.

## Data Availability

Data is available upon request from the National Emergency Medical Service Information System

https://nemsis.org/using-ems-data/request-research-data/

## Acknowledgment

The authors appreciate the National Highway Traffic Safety Administration’s Office of Emergency Medical Service and the Technical Assistance Center at the University of Utah for providing the data.

## References

1. Centers for Disease Control and Prevention. Older Adult Drivers 2021 [12/13/2021]. Available from: https://www.cdc.gov/transportationsafety/older_adult_drivers/index.html

2. National Institute for Occupational Safety and Health. Older Drivers in the Workplace: How Employers and Workers Can Prevent Crashes: Centers for Disease Control and Prevention; 2016 [04/15/2022]. Available from: https://www.cdc.gov/niosh/docs/2016-116/pdfs/2016-116.pdf

3. Federal Highway Administration. Nighttime Visibility: United States Department of Transportation; 2022 [03/08/2023]. Available from: https://safety.fhwa.dot.gov/roadway_dept/night_visib/general-information.cfm

4. Centers for Disease Control and Prevention, National Center for Injury Prevention and Control. Overall All Transport Nonfatal Emergency Department Visits and Rates per 100,000: 2019, United States, All Races, Both Sexes, Ages 65 to 85+, Disposition: All Cases Atlanta, GA: CDC; 2022 [04/15/2022]. Available from: https://wisqars.cdc.gov/nonfatal-reports

5. Centers for Disease Control and Prevention, National Center for Injury Prevention and Control. Overall MV-Occupant Nonfatal Emergency Department Visits and Rates per 100,000: 2019, United States, All Races, Both Sexes, Ages 65 to 85+, Disposition: All Cases Atlanta, GA: CDC; 2022 [04/15/2022]. Available from: https://wisqars.cdc.gov/nonfatal-reports

6. Centers for Disease Control and Prevention, National Center for Injury Prevention and Control. Overall Pedestrian Nonfatal Emergency Department Visits and Rates per 100,000, 2009 - 2020, United States: All Races, Both Sexes, Ages 65 to 85+, Disposition: All Cases Atlanta, GA: CDC; 2022 [04/15/2022]. Available from: https://wisqars.cdc.gov/nonfatal-reports

7. Alcañiz M, Santolino M, Ramon L. Drinking patterns and drunk-driving behaviour in Catalonia, Spain: A comparative study. Transportation Research Part F: Traffic Psychology and Behaviour. 2016;42:522–531.

8. Bondallaz P, Favrat B, Chtioui H, et al. Cannabis and its effects on driving skills. Forensic Science International. 2016;268:92–102.

9. Kumar S, Bansal YS, Singh D, et al. Alcohol and Drug Use in Injured Drivers - An Emergency Room Study in a Regional Tertiary Care Centre of North West India. Journal of Clinical and Diagnostic Research. 2015 Jul;9(7):Hc01–4.

10. Freeman DG. Drunk driving legislation and traffic fatalities: New evidence on BAC 08 laws. 2007;25(3):293–308.

11. Hoots BE, Xu L, Kariisa M, et al. 2018 Annual surveillance report of drug-related risks and outcomes--United States 2018 [cited.

12. Mattson M, Lipari RN, Hays C, et al. A day in the life of older adults: Substance use facts. The CBHSQ report. 2017 [cited. https://www.ncbi.nlm.nih.gov/books/NBK436750/pdf/Bookshelf_NBK436750.pdf

13. DiMaggio CJ, Avraham JB, Frangos SG, et al. The role of alcohol and other drugs on emergency department traumatic injury mortality in the United States. Drug and Alcohol Dependence. 2021 2021/08/01/;225:108763.

14. Adeyemi OJ, Paul R, DiMaggio CJ, et al. An assessment of the non-fatal crash risks associated with substance use during rush and non-rush hour periods in the United States. Drug and Alcohol Dependence. 2022 2022/03/03/:109386.

15. Asefa NG, Ingale L, Shumey A, et al. Prevalence and factors associated with road traffic crash among taxi drivers in Mekelle town, northern Ethiopia, 2014: a cross sectional study. PLoS One. 2015;10(3):e0118675.

16. Compton RP, Berning A. Drug and Alcohol Crash Risk. Traffic Safety Facts: Research Note. 2015 [cited. Behavioral Safety Research available at http://www.nhtsa.gov/staticfiles/nti/pdf/812117-Drug_and_Alcohol_Crash_Risk.pdf

17. Penmetsa P, Pulugurtha SS. Risk drivers pose to themselves and other drivers by violating traffic rules. Traffic Injury Prevention. 2017;18(1):63–69.

18. Blows S, Ivers RQ, Connor J, et al. Marijuana use and car crash injury. Addiction. 2005;100(5):605–11.

19. Preuss UW, Huestis MA, Schneider M, et al. Cannabis Use and Car Crashes: A Review [Systematic Review]. Frontiers in psychiatry. 2021 2021-May-28;12.

20. Brubacher JR, Chan H, Erdelyi S, et al. Cannabis use as a risk factor for causing motor vehicle crashes: a prospective study. Addiction. 2019 Sep;114(9):1616–1626.

21. Drummer OH, Gerostamoulos D, Di Rago M, et al. Odds of culpability associated with use of impairing drugs in injured drivers in Victoria, Australia. Accid Anal Prev. 2020 Feb;135:105389.

22. Drummer OH, Gerostamoulos J, Batziris H, et al. The involvement of drugs in drivers of motor vehicles killed in Australian road traffic crashes. Accident Analysis & Prevention. 2004 2004/03/01/;36(2):239–248.

23. Laumon B, Gadegbeku B, Martin JL, et al. Cannabis intoxication and fatal road crashes in France: population based case-control study. Bmj. 2005 Dec 10;331(7529):1371.

24. Byrne JP, Mann NC, Dai M, et al. Association Between Emergency Medical Service Response Time and Motor Vehicle Crash Mortality in the United States. JAMA surgery. 2019;154(4):286–293.

25. Adeyemi OJ, Paul R, Arif A. An assessment of the rural-urban differences in the crash response time and county-level crash fatalities in the United States. The Journal of Rural Health. 2021 2021/10/19.

26. Adeyemi OJ, Paul R, DiMaggio C, et al. The association of crash response times and deaths at the crash scene: A cross-sectional analysis using the 2019 National Emergency Medical Service Information System. The Journal of Rural Health. 2022 2022/04/22.

27. Cabrera-Arnau C, Prieto Curiel R, Bishop SR. Uncovering the behaviour of road accidents in urban areas. Royal Society open science. 2020 Apr;7(4):191739.

28. Insurance Institute for Highway Safety. Fatality Facts 2019: Urban/rural comparison: Insurance Institute for Highway Safety, Highway Loss Data Institute; 2022 [04/15/2022]. Available from: https://www.iihs.org/topics/fatality-statistics/detail/urban-rural-comparison

29. Rural Health Information Hub. Substance Use and Misuse in Rural Areas 2020 [04/15/2022]. Available from: https://www.ruralhealthinfo.org/topics/substance-use

30. Vespa J. The U.S. Joins Other Countries With Large Aging Populations: United States Census Bureau; 2021 [04/15/2022]. Available from: https://www.census.gov/library/stories/2018/03/graying-america.html

31. Pallin DJ, Espinola JA, Camargo CA, Jr. US population aging and demand for inpatient services. J Hosp Med. 2014 Mar;9(3):193–6.

32. National Emergency Medical Services Information System. How NEMSIS Works 2019 [11/27/2020]. Available from: https://nemsis.org/what-is-nemsis/how-nemsis-works/

33. NEMSIS. Research Data Resources 2023 [03/09/2023]. Available from: https://nemsis.org/using-ems-data/request-research-data/research-data-resources/

34. Schreiner B, Salter J. Kentucky hardest hit as storms leave dozens dead in 5 states. Chattanooga Times Free Press. 2021 December 11, 2021 [cited 03/12/2023]. Available from: https://www.timesfreepress.com/news/2021/dec/11/2-dead-tennessee-severe-storms-move-through-state-/

35. Childs JW. Newnan, Georgia, Tornado: ’Our Hearts Are Broken’. The Weather Channel. 2021 [cited 03/12/2023]. Available from:https://weather.com/news/news/2021-03-26-severe-weather-outbreak-impacts-alabama-mississippi-tennessee-georgia

36. National Weather Service. Tornadoes of March 3, 2019: Event Summary for Central Alabama 2019 [11/08/2021]. Available from: https://www.weather.gov/bmx/event_03032019

37. Emergency Medical Services. NEMSIS Data Dictionary2020 [cited. https://nemsis.org/media/nemsis_v3/release-3.4.0/DataDictionary/PDFHTML/DEMEMS/index.html

38. Beeson MS, Ankel F, Bhat R, et al. The 2019 Model of the Clinical Practice of Emergency Medicine. Journal of Emergency Medicine. 2020;591(1):96–120.

39. Counselman FL, Babu K, Edens MA, et al. The 2016 Model of the Clinical Practice of Emergency Medicine. The Journal of emergency medicine. 2017 Jun;52(6):846–849.

40. National Emergency Medical Services Information System. eHistory.17 -Alcohol/Drug Use Indicators: National Emergency Medical Services Information System; 2021 [04/14/2022]. Available from: https://nemsis.org/media/nemsis_v3/release-3.4.0/DataDictionary/PDFHTML/DEMEMS/sections/elements/eHistory.17.xml

41. National Emergency Medical Services Information System. Defined Lists and Other Resources 2021 [05/10/2022]. Available from: https://nemsis.org/technical-resources/version-3/version-3-resources/

42. Adeyemi OJ, Arif AA, Paul R. Exploring the relationship of rush hour period and fatal and non-fatal crash injuries in the U.S.: A systematic review and meta-analysis. Accid Anal Prev. 2021 Oct 28;163:106462.

43. Lee KJ, Carlin JB. Multiple imputation for missing data: fully conditional specification versus multivariate normal imputation. Am J Epidemiol. 2010 Mar 1;171(5):624–32.

44. NEMSIS Technical Assistance Center. National EMS Database NEMSIS Public Release Research Data Set: 2020 User Manual2021 [cited. https://nemsis.org/wp-content/uploads/2021/05/2020-NEMSIS-RDS-340-User-Manual_v3-FINAL.pdf

45. Dray S, Josse J. Principal component analysis with missing values: a comparative survey of methods. Plant Ecology. 2015;216(5):657–667.

46. McNeish D. Exploratory Factor Analysis With Small Samples and Missing Data. J Pers Assess. 2017 Nov-Dec;99(6):637–652.

47. Williams R. Gologit2: A program for generalized logistic regression/partial proportional odds models for ordinal variables. STATA Journal. 2005;12:2005.

48. SAS Institute Inc. SAS 9.4. 9.4. Cary, NC: SAS Institute Inc; 2019.

49. StataCorp. Stata Statistical Software: Release 17. College Station, TX: StataCorp LLC; 2020.

50. de Jong MR, Van der Elst M, Hartholt KA. Drug-related falls in older patients: implicated drugs, consequences, and possible prevention strategies. Ther Adv Drug Saf. 2013 Aug;4(4):147–54.

51. Shakya I, Bergen G, Haddad YK, et al. Fall-related emergency department visits involving alcohol among older adults. Journal of safety research. 2020;74:125–131.

52. Sun Y, Zhang B, Yao Q, et al. Association between usual alcohol consumption and risk of falls in middle-aged and older Chinese adults. BMC Geriatr. 2022 2022/09/14;22(1):750.

53. Moore AR, O’Keeffe ST. Drug-induced cognitive impairment in the elderly.Drugs Aging. 1999 Jul;15(1):15–28.

54. Zhang R, Shen L, Miles T, et al. Association of Low to Moderate Alcohol Drinking With Cognitive Functions From Middle to Older Age Among US Adults. JAMA Network Open. 2020;3(6):e207922–e207922.

55. Moore AA, Whiteman EJ, Ward KT. Risks of combined alcohol/medication use in older adults. The American Journal of Geriatric Pharmacotherapy. 2007 2007/03/01/;5(1):64–74.

56. Narkiewicz K, Cooley RL, Somers VK. Alcohol Potentiates Orthostatic Hypotension. Circulation. 2000 2000/02/01;101(4):398–402.

57. Husain K, Ansari RA, Ferder L. Alcohol-induced hypertension: Mechanism and prevention. World J Cardiol. 2014;6(5):245–252.

58. Klatsky AL, Gunderson E. Alcohol and hypertension: a review. J Am Soc Hypertens. 2008 Sep-Oct;2(5):307–17.

59. Smink BE, Egberts ACG, Lusthof KJ, et al. The relationship between benzodiazepine use and traffic accidents: A systematic literature review. CNS Drugs. 2010;24(8):639–653.

60. Li G, Chihuri S. Prescription opioids, alcohol and fatal motor vehicle crashes: a population-based case-control study. Injury epidemiology. 2019;6:11–11.

61. Dixon MA, Chartier KG. Alcohol Use Patterns Among Urban and Rural Residents: Demographic and Social Influences. Alcohol Res. 2016;38(1):69–77.

62. Derefinko KJ, Bursac Z, Mejia MG, et al. Rural and urban substance use differences: Effects of the transition to college. The American journal of drug and alcohol abuse. 2018;44(2):224–234.

63. Governors Highway Safety Association. America’s rural roads: Beautiful and deadly2022 [cited. https://www.ghsa.org/sites/default/files/2022-09/America%E2%80%99s%20Rural%20Roads%20-%20Beautiful%20and%20Deadly%20FNL.pdf

64. Adeyemi O, Paul R, Arif A. Spatial Cluster Analysis of Fatal Road Accidents From Non-Use of Seat Belts Among Older Drivers. Innovation in Aging. 2020;4(Supplement_1):113–114.

65. Adeyemi OJ, Paul R, DiMaggio C, et al. Rush Hour-Related Road Crashes: Assessing the Social and Environmental Determinants of Fatal and Non-Fatal Road Crash Events [Ph.D.]. Ann Arbor: The University of North Carolina at Charlotte; 2021.

66. Insurance Institute for Highway Safety. Fatality Facts 2018: Urban/rural comparison: Insurance Institute for Highway Safety/Highway Loss Data Institute; 2019 [12/5/2020]. Available from: https://www.iihs.org/topics/fatality-statistics/detail/urban-rural-comparison

67. National Center for Statistics and Analysis. Rural/Urban Comparison of Traffic Fatalities. Traffic Safety Fact: 2017 Data. 2019 [cited. https://crashstats.nhtsa.dot.gov/Api/Public/ViewPublication/812741

68. Prochaska JO, Redding CA, Evers KE. The transtheoretical model and stages of change. Health Behavior: Theory, Research, and Practice. 2015;97.

69. Watakakosol R, Suttiwan P, Ngamake ST, et al. Integration of the Theory of Planned Behavior and Transtheoretical Model of Change for Prediction of Intentions to Reduce or Stop Alcohol Use among Thai Adolescents. Subst Use Misuse. 2021;56(1):72–80.

70. Berning A, Smither DD. Understanding the limitations of drug test information, reporting, and testing practices in fatal crashes: traffic safety facts: research note. United States. Department of Transportation. National Highway Traffic Safety …; 2014.

71. DiMaggio C, Wheeler-Martin K, Oliver J. Alcohol-Impaired Driving in the United States: Review of Data Sources and Analyses. 2018 08/06/2021. In: Getting to Zero Alcohol-Impaired Driving Fatalities: A Comprehensive Approach to a Persistent Problem [Internet]. The National Academies Press. Available from: https://www.ncbi.nlm.nih.gov/books/NBK500064/.

